# Biochemical analyses can complement sequencing-based ARG load monitoring: a case study in Indian hospital sewage networks

**DOI:** 10.1101/2024.05.31.24308262

**Authors:** S. Bhanushali, K. Pärnänen, D. Mongad, D. Dhotre, L. Lahti

**Author notes:** These authors contributed equally as first authors (S. Bhanushali, K. Pärnänen). Equal contribution (S. Bhanushali, K. Pärnänen, D. Dhotre, L. Lahti).

## Abstract

Antibiotic resistance is an emerging global crisis which has been estimated to cause increasing numbers of deaths. Low and middle-income countries (LMICs) are challenged with a larger burden of antibiotic resistance, as antibiotic resistance is more common in LMICs, and access to antibiotics and health care is often limited compared to high-income countries. Further exacerbating the issue is the possible lack of efficient treatment of hospital sewage which can have high concentrations of clinically relevant antibiotic-resistant pathogens. Monitoring of antibiotic resistance genes (ARGs) in sewage along the sewage networks (from hospitals to community sewers and sewage treatment plant effluents) would provide crucial tools for identifying hotspots of ARG pollution. However, the methods that are currently used to quantify ARGs rely on expensive shotgun sequencing or qPCR. Therefore, we investigated whether ARG load monitoring could be complemented with inexpensive standard biochemical analyses. Our results show that across four different sewage networks and three seasons, biological oxygen demand (BOD) and total organic carbon (TOC) can provide robust indicators of total ARG load. This lays grounds for finding cost-efficient techniques for sewage ARG pollution monitoring in low-resource settings.

## Introduction

Antimicrobial resistance (AMR) has become a critical health challenge, threatening decades of medical progress by complicating the clinical protocols and ultimately increasing healthcare costs. With its rapid spread, AMR is not only confined to clinical settings but has become prevalent across natural ecosystems, agricultural settings, and urban infrastructures, covering the three key elements of One Health (Lebov et al., 2017, Karkman et al., 2019).

The development and spread of AMR is a multifactorial phenomenon (Pärnänen et al., al 2019, Nadimpalli et al., 2020), where hospital-borne waste and community sewage have emerged as a significant concern. For instance, the AWARE (Antibiotic Resistance in Wastewater) study highlights the transmission risks posed by hospital wastewater to local communities and Sewage Treatment Plant (STP) employees (Wengenroth et al., 2021). Hospital sewage is a hotspot for antimicrobial-resistant bacteria, including the ESKAPE category pathogens viz. *Enterococcus faecium, Staphylococcus aureus, Klebsiella pneumoniae, Acinetobacter baumannii, Pseudomonas aeruginosa, and Enterobacter spp*. (Denissen et al., 2022) These pathogens (also on the WHO priority list, Davies et al., 2017), beyond their infectivity, are now considered to be environmental contaminants along with the ARGs they host.

Community sewers and STPs play an important role in AMR transmission especially if they receive untreated hospital sewage. However, STPs typically reduce the relative abundance of ARGs in the influent sewage when they function as intended (Pärnänen et al., 2019) resulting in a lower relative abundance of ARGs in the effluents. However, the STPs in many Low and Middle-Income Countries such as Sri Lanka, have been reported to promote resistance (Kumar et al., 2020).

Malfunctioning sewage treatment poses a major threat to human health, and monitoring the ARG reduction capability in addition to the release of pathogens is needed to be able to intervene with problematic treatment sites. However, ARG monitoring using molecular techniques such as qPCR or shotgun sequencing is not feasible due to a lack of resources. Thus, methods which can be readily implemented with lower resources are needed to complement these techniques. Complements to molecular biology techniques could include enumerating coliforms, or using biochemical analyses such as biological oxygen demand (BOD) or total organic carbon (TOC). In this study, we investigated if BOD or TOC could be used to estimate the ARG loads in sewage. We sampled four hospitals and their downstream sewage (hospital and community sewage, STP influent and STP effluent) in three seasons (spring, autumn, and monsoon) to account for variability in ARGs and bacterial taxa in different sites and seasons.

## 2 Sample origin and collection

### 2.1. Site selection

The study was conducted in an Indian metropolis area with a high population density, representing a significant demographic, economic, and cultural diversity (World Urban Areas-Demographia 2018). Sampling permission was granted by the local governing authorities and all the necessary precautionary measures were undertaken while sampling. We selected four different locations for sampling, pairing four Sewage Treatment Plants (STPs) with four local district hospitals. The locations were selected in a way that major administrative divisions of the city are represented. Site W is a part of South-zone, site B East-zone, site M North-Zone, and site G is undefined.

The hospitals selected offer general and multi-speciality services with an average capacity of 470 beds per hospital covering a broad range of in-patient and out-patient treatment alternatives likely to influence waste generation and thus the resistome determinants and microbiome features in the sewage. For instance, in developing or semi-developed countries, the average wastewater generated by hospitals is around 290 m^3^/hospital/day and 250 L/patient/day (Majumder et al., 2021). Within each hospital, the sampling point represented the entire hospital’s sewage. The sampling points for all the hospitals were located within the premises (in the range of ∼50-80 metres) but outside the main building. For community sewage, the access points or manhole covers represented the residential areas. Both the hospital and community effluents are untreated before discharge into the main sewage network and sampled via access points/manhole covers along the network ending in STPs. The city generates ∼3000 Million Litres per Day (MLD) of sewage; of which ∼2100 MLD gets treated.

Considering the current operational capacity of the STPs we selected, over half of the total treated sewage is handled by the four STPs alone. On average, each STP treats ∼320 MLD sewage at the 1° (primary) level of treatment. The sewage entering the STPs is a mixture of urban sewage originating from the local residential and commercial regions (e.g. hospitals in this case) and is treated before being discharged. Sample collection at the influent and effluent sampling points of STPs was done using the same method except that it was performed independently of the manholes.

### 2.2 Sample collection and processing

Grab sampling was performed across three seasons: Monsoon (August 2019), Autumn (October 2019), and Spring (February 2020) at four different locations described earlier to account for spatio-temporal variations. The daily mean maximum temperature during the sampling months is 30.2°C (August), 33.6°C (October) and 31.7°C (February) [Indian Meteorological Department, Pune]. Figure 1 provides a schematic illustration of the sampling locations and sampling points along the urban wastewater network.

**Figure 1.**
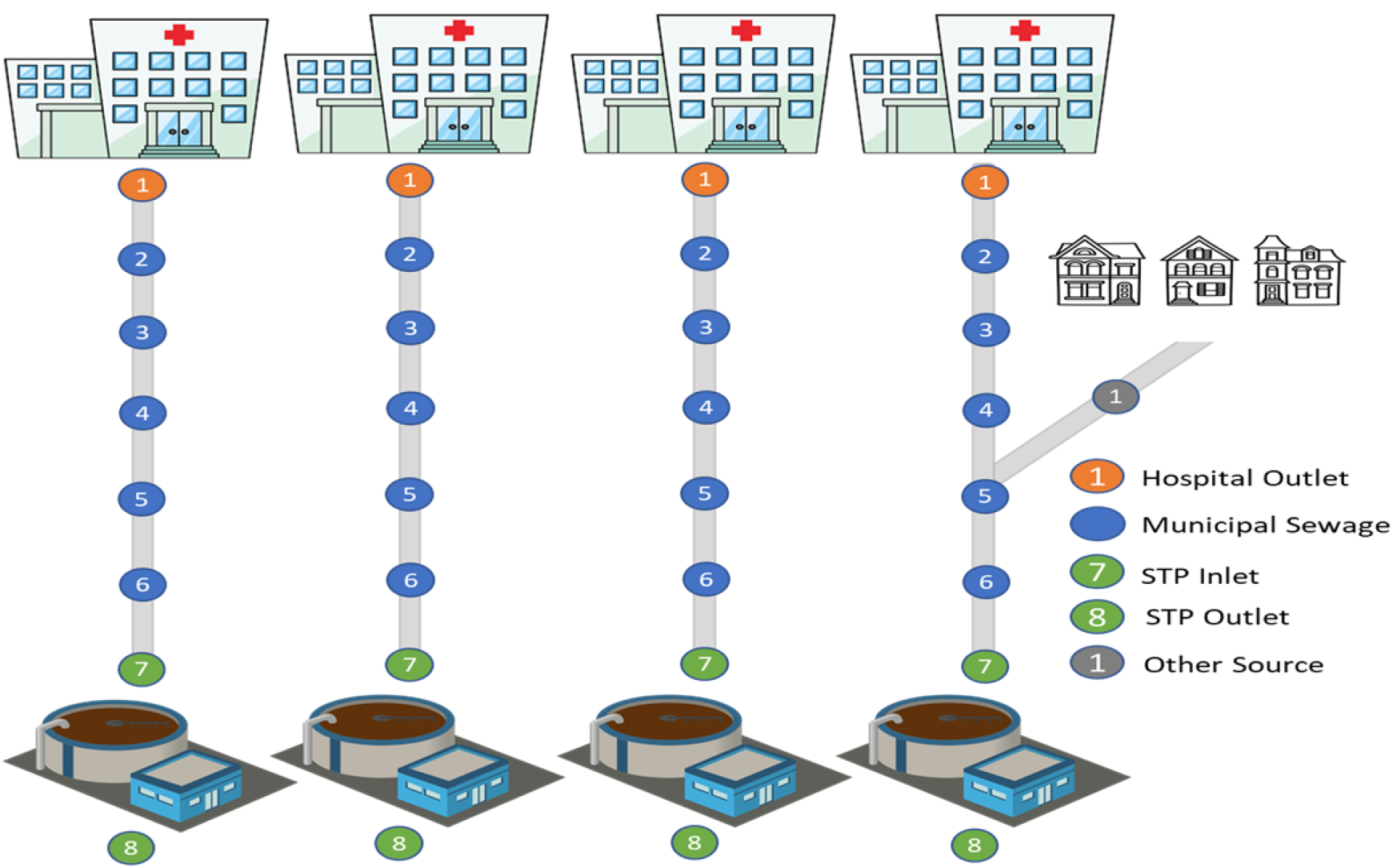
Schematic representation of the process flow involving hospitals, residential areas and STPs. Each vertical sequence represents a distinct pathway from hospitals (top) through various points (1 to 8) to the STPs (bottom). The sub-sequence includes an additional residential area component, illustrating an alternative pathway in the system.

For each location, four samples were collected: Hospital outlet (point 1), Community sewer (point 4), STP inlet (point 7), and STP outlet (point 8). A total of 48 samples were collected across four locations and three seasons. Approximately 2 litres of sewage samples from each sampling point were collected in clean, ultraviolet light-disinfected plastic containers and transported to the laboratory in a cold chain maintained at 4°C and preserved until further processing. Bio-physicochemical characterization of sewage samples was performed both onsite and offsite. Onsite analysis was performed using a portable meter (Hanna HI-98194, Hanna Instruments Limited) to yield data on sample pH, dissolved oxygen(%), temperature(°C), conductivity(µS/cm), and Total Dissolved Solids (TDS). Off-site sample processing was performed using an in-house benchtop photometer (HI-83300-02, Hanna Instruments Limited) to record Iron, Manganese and Phosphate levels. Additionally, standard parameters: Total Suspended Solids (TSS), Total Solids (TS), Biological Oxygen Demand (BOD), Chemical Oxygen Demand (COD), Total Organic Carbon (TOC), Sulphur and Nitrogen were analysed.

The parameters are recorded in mg/L unless otherwise stated. All the analytical procedures were performed by following the manufacturer’s instructions where relevant by using appropriate standards.

### 2.3 Metagenomic DNA Extraction and Next-Generation Sequencing

A 250 mL aliquot from each sewage sample was centrifuged, the supernatants were discarded, and the resulting pellets (approximately 1-1.5 g) were suspended in lysis buffer (QIAGEN N.V.), and treated with lysozyme (100 mg/mL) and proteinase K (0.2 mg/mL) (Thermo Scientific, USA). Total DNA was extracted from the pre-treated sample pellet using the QIAamp Fast DNA Stool Mini Kit (QIAGEN N.V.) following the manufacturer’s instructions. The eluted DNA concentration and purity were recorded using a NanoDrop 2000 spectrophotometer and a Qubit 2.0 fluorometer and further, the samples were stored at ultra-low temperature. High-quality metagenomic DNA samples were sequenced on the Illumina NovaSeq-6000 platform using 150×2 sequencing chemistry.

### 2.4 Bioinformatic and statistical analysis

All quality control analysis and read mapping were executed using a custom in-house Snakemake workflow (v8.11.5) [Mölder et al., 2021]. Raw-read processing was performed in the computing infrastructure service provided by CSC-IT Center for Science, Finland. On average ∼29 million raw reads per sample generated were processed through the workflow’s quality control step which includes a MultiQC (v.1.9) [Ewels et al., 2016] assessment to inspect low-quality reads. Following this a(one) sample not satisfying the quality criteria was omitted from the downstream analysis. Eventually, high-quality nucleotide sequence reads from 47 samples were mapped using Bowtie2 (v2.5.3) (-D 20 -R 3 -N 1 -L 20 -i S,1,0.5) [Langmead & Salzberg 2012] against the ResFinder database (v.2.1.1) [Gschwind et al., 2023] to identify reads that map to ARGs. MetaPhlAn (v4.1.1) [Míguez et al., 2023] was used for generating taxonomic features. Counts from the ResFinder output were normalised by the Reads Per Kilobase Million (RPKM) method to account for the variations arising from differences in gene lengths and sequencing depth/coverage. RPKM normalised counts were further used for statistical testing and visualisation. The ARG load per sample was calculated by summing the RPKM values of individual genes in each sample. Statistical testing and visualisation were done using R (v4.3.3) and a combination of packages; phyloseq (v1.46.0) [McMurdie et al., 2013], ggplot2 (v3.5.0), dplyr (v.1.4), vegan (v2.6-4), RColorBrewer (v1.1-3), viridis (v0.6.5).

## 3 Results

### 3.1 Community composition

First, we characterized how bacterial community composition in the sewage water is impacted by the relevant background factors including season, hospital district, and sampling site. The taxonomic composition varied according to season and hospital district but not by sampling site (Bray-Curtis dissimilarity; PERMANOVA, season: R^2^ = 0.18, *p* = 0.001; hospital district: R^2^ = 0.12, *p* = 0.003, sampling point R^2^ = 0.7, *p* = 0.155). Samples taken during monsoon had distinct taxonomic composition from the spring and autumn seasons and separated from the other seasons along the first axis of the PCoA (Figure 4). More specifically, during monsoon season the relative abundance of *Pseudomonas* was higher while *Streptococcus* lower abundance (Figure 2). Interestingly, the sampling point was not a significant determinant of the species and even the furthest sampling point in the system STP effluent was not significantly different from the hospital sewage (pairwise adonis, Bray-Curtis, R^2^ = 0.08, *p* = 0.08). However, the hospital districts had sometimes significantly different species (pairwise adonis, Bray-Curtis, B-M, G-M and G-W, R^2^ = 0.096-0.12, FDR<0.05) In general, the sewage contained high abundances of bacterial genera typical for the human gut as well as potential pathogenic taxa whose resistant members are part of ESKAPE.

**Figure 2.**
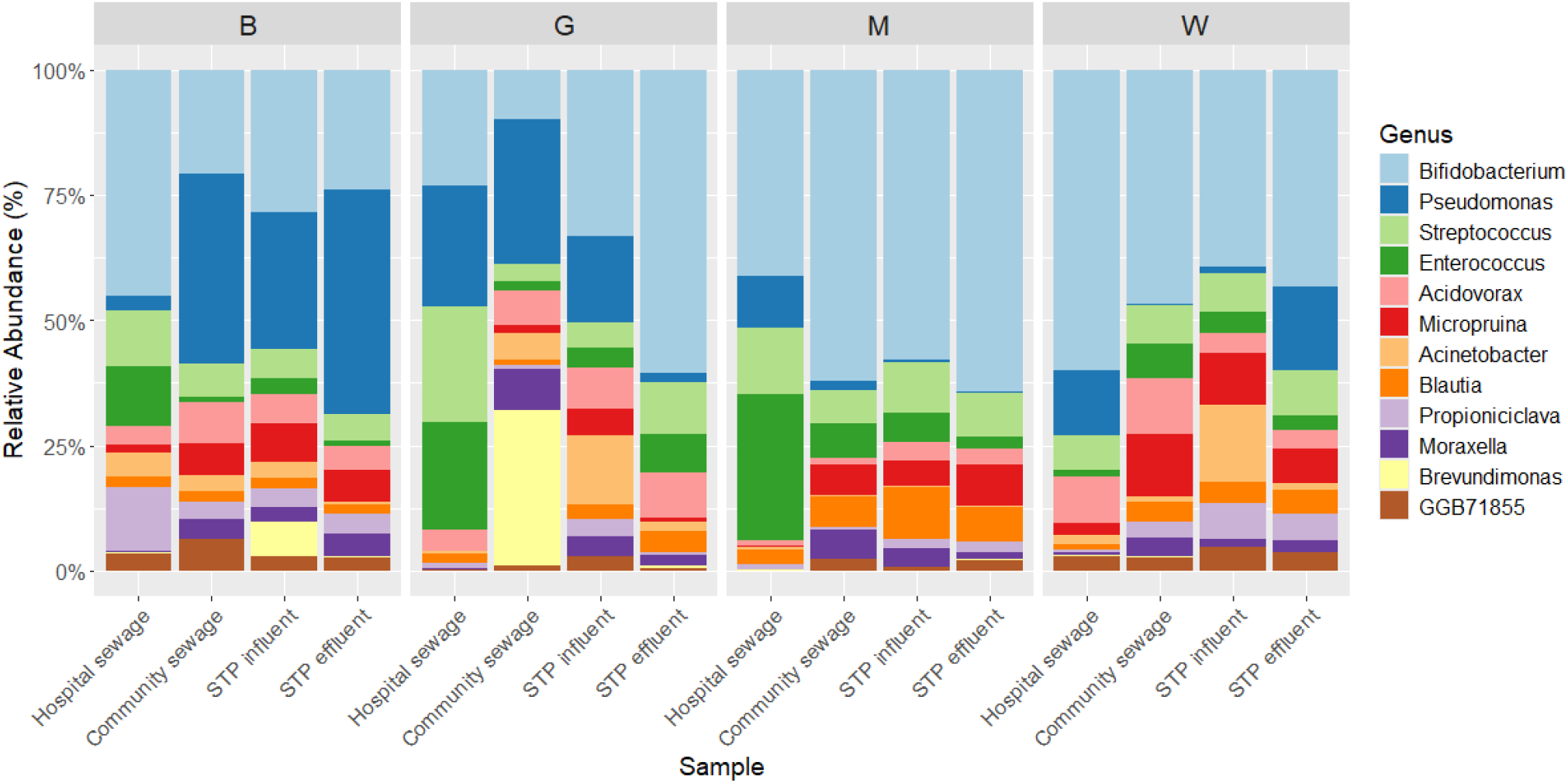
Community composition. Average genus-level community composition (y-axis) for the most dominant genera across the different sample types (bars) and locations (individual panels). The genera are arranged in descending order based on their relative abundance.

**Figure 3.**
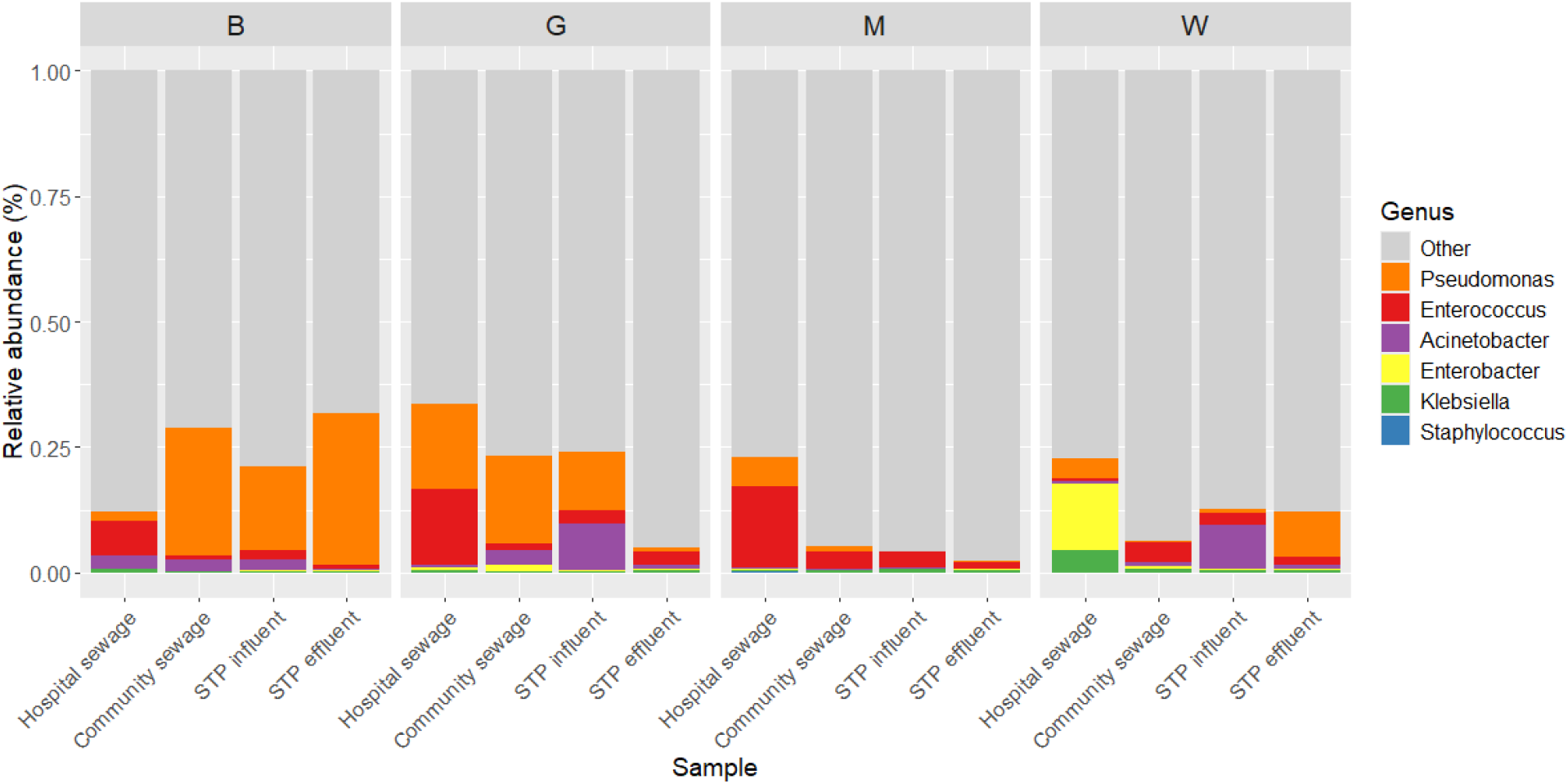
Relative abundance of genera belonging to ESKAPE pathogens. Stacked bar plot showing relative composition (y-axis) of ESKAPE genera across different sample types along the x-axis with samples categorized by locations (individual panels).

**Figure 4.**
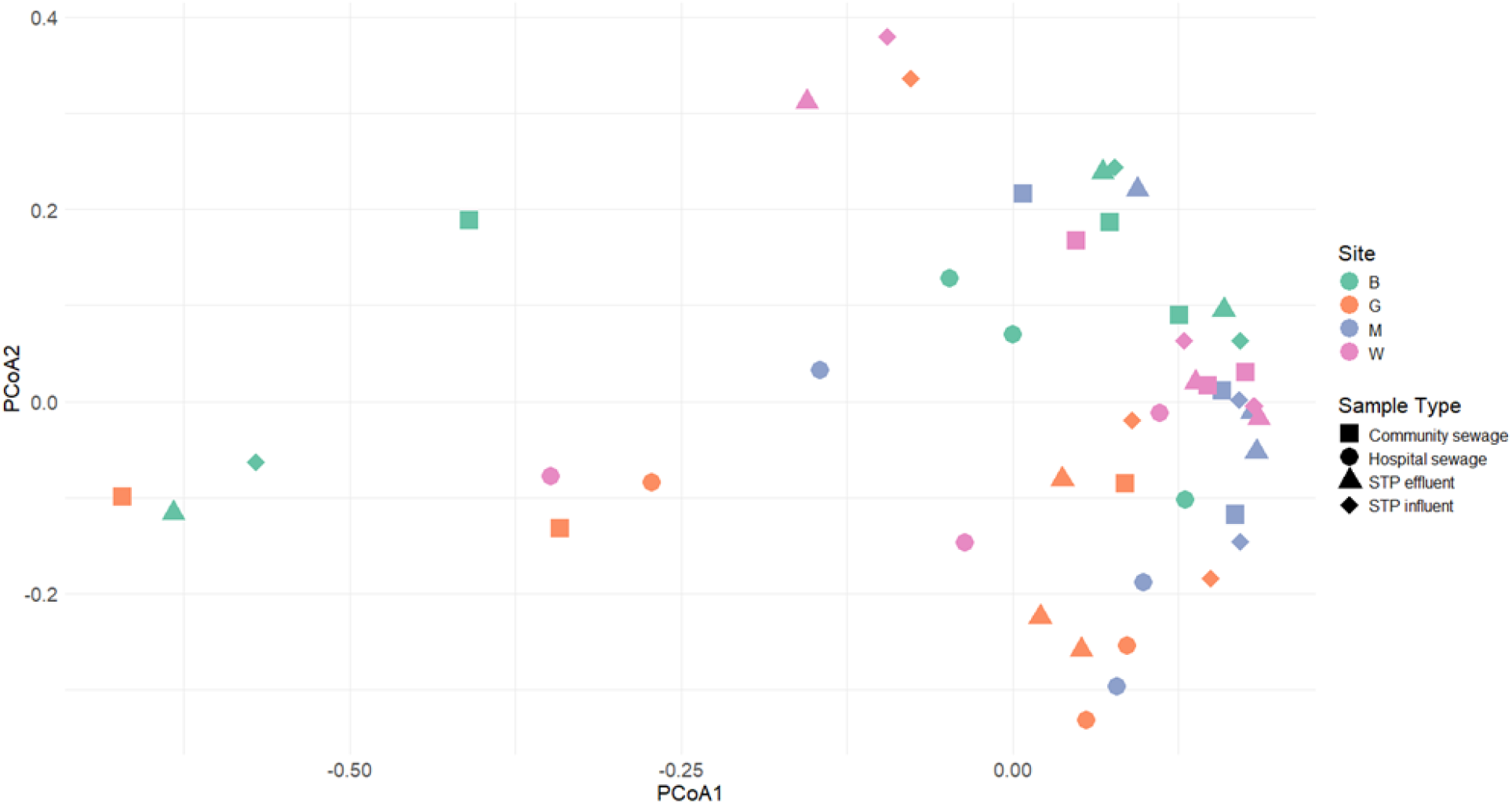
Variation in species-level bacterial community composition. Principal Coordinates Analysis of sewage samples (PCoA; species-level relative abundance; **Bray-Curtis dissimilarity)** Sampling location is indicated by color, and sample type by shape.

**Figure 5.**
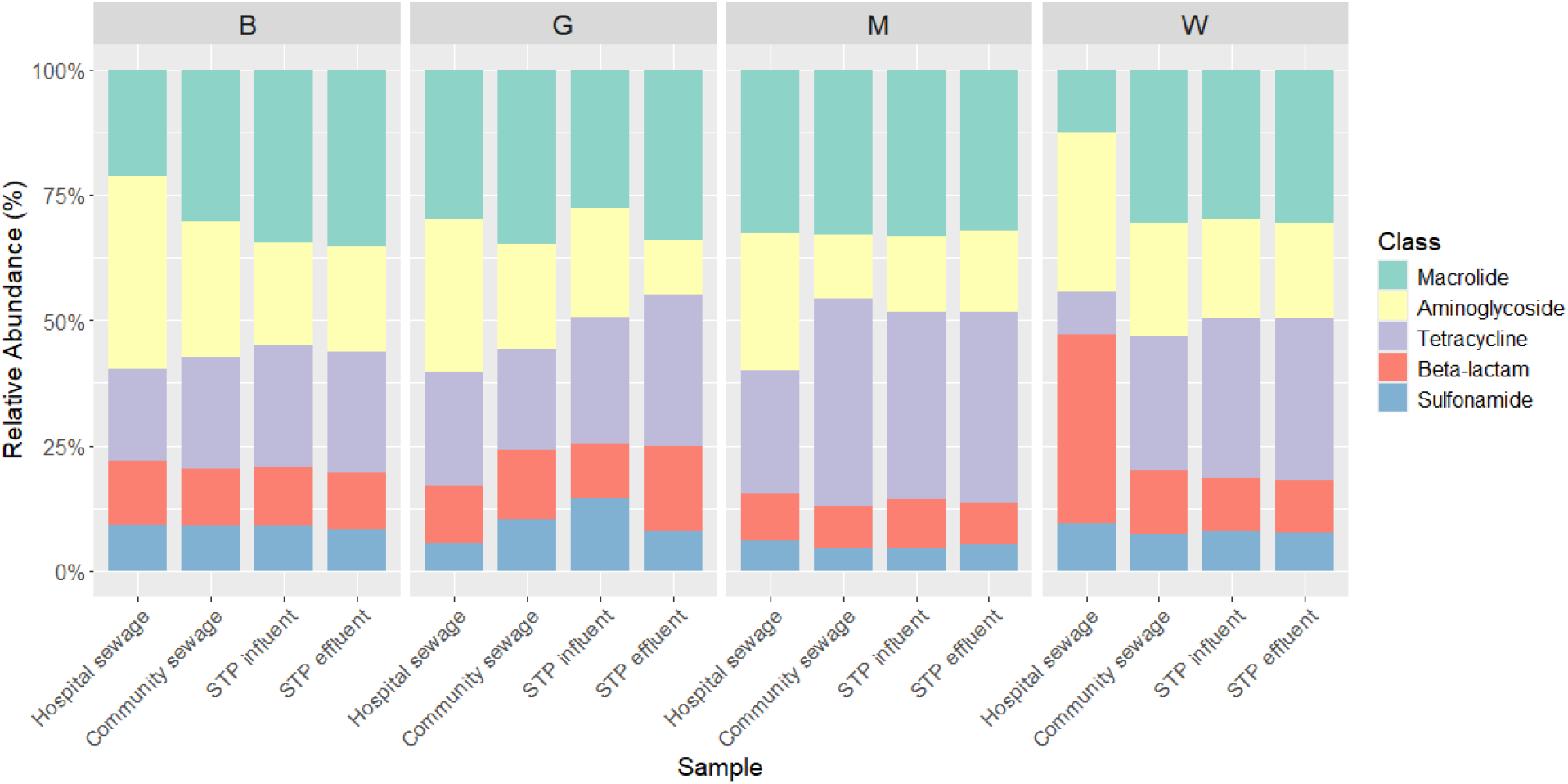
Average relative abundance of antibiotic resistance gene classes. Most abundant antibiotic resistance gene classes among the sample types (x-axis) and locations (panels).

**Figure 6.**
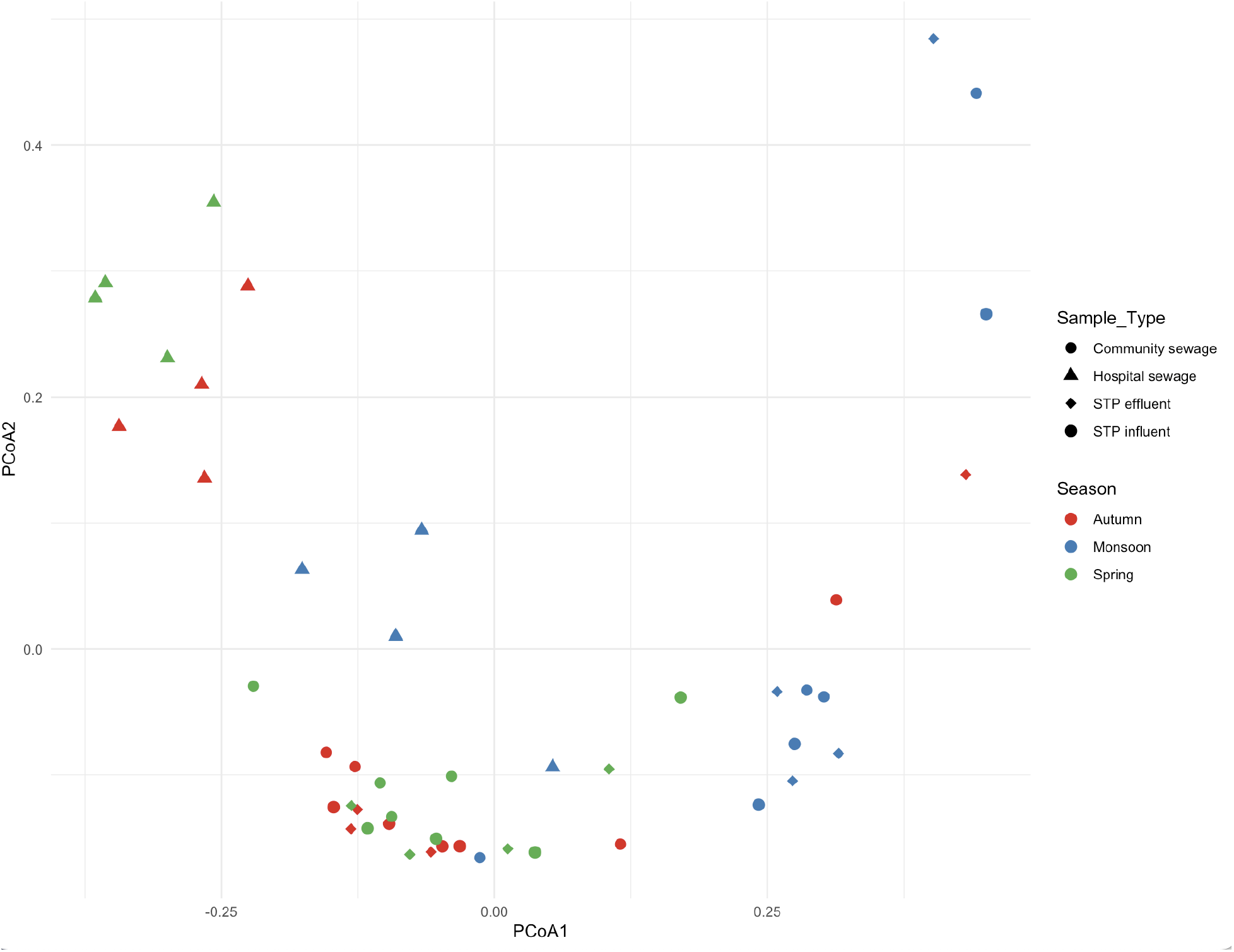
Variation in resistome profiles. Principal Coordinates Analysis (PCoA; of the ARG profiles; Bray-Curtis dissimilarity). The colors indicate sampling locations while the shapes denote the sample types.

### 3.2 Resistome variation

Resistome variation (beta diversity of the ARG profiles) was significantly associated with the season, site and sampling point (adonis, Bray-Curtis, sampling point: R^2^ = 0.18, *p* = 0.001, season: R^2^ = 0.144, *p* = 0.001, hospital district: R^2^ = 0.11, *p* = 0.001). The resistome composition of hospital sewage samples differed from the other sampling points (R^2^ = 0.18-0.24, FDR=0.002) with higher relative abundance of aminoglycoside and beta-lactam resistance genes as compared to the other sewage samples, and lower abundance of tetracycline resistance genes. The seasonal variance could also be observed between monsoon and the other seasons (R^2^ 0.12-0.16, FDR = 0.0015), the relative abundance of tetracycline resistance genes was higher during monsoon.

### 3.3 ARG load

The ARG load was highest in the hospital sewage. Community sewage and STP influent and effluent had similar ARG loads. Notably, there was no clear trend that the ARG load would decrease from STP influent to effluent. The ARG load was lower in the monsoon season, possibly because increased rainfall causes more environmental bacteria to flow into the sewage system.

### 3.4 Association between bio-physiochemical measurements and ARG load

BOD was positively associated with the ARG load in the sewage and STP influents and effluents (linear model, estimate = x 95% CI x, p = x, n = 47) the trend was positive in all sample types and seasons Figure 9. The change in ARG load from influent to effluent with the respective change in BOD was not significantly associated with each other (linear model, adjusting for hospital district and season, estimate = x, 95% CI -x, p = x, n= x). Thus, the result is inconclusive for whether BOD can be used to predict the efficacy of STP treatment in reducing the ARG load.

**Figure 7.**
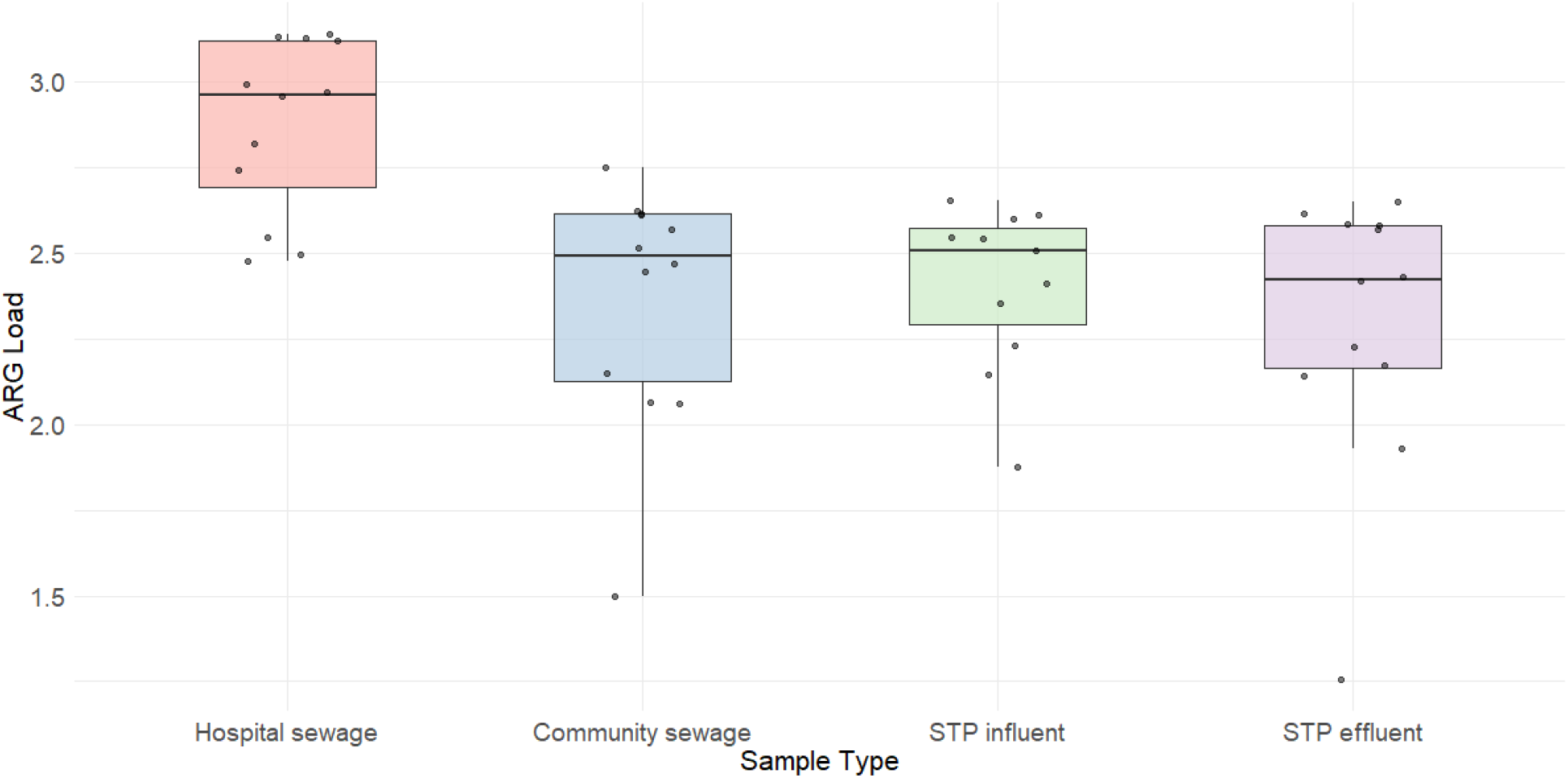
Median distribution of total ARG load (y-axis) detected among the sample types (x-axis) with the jitters representing the samples within each group.

**Figure 8.**
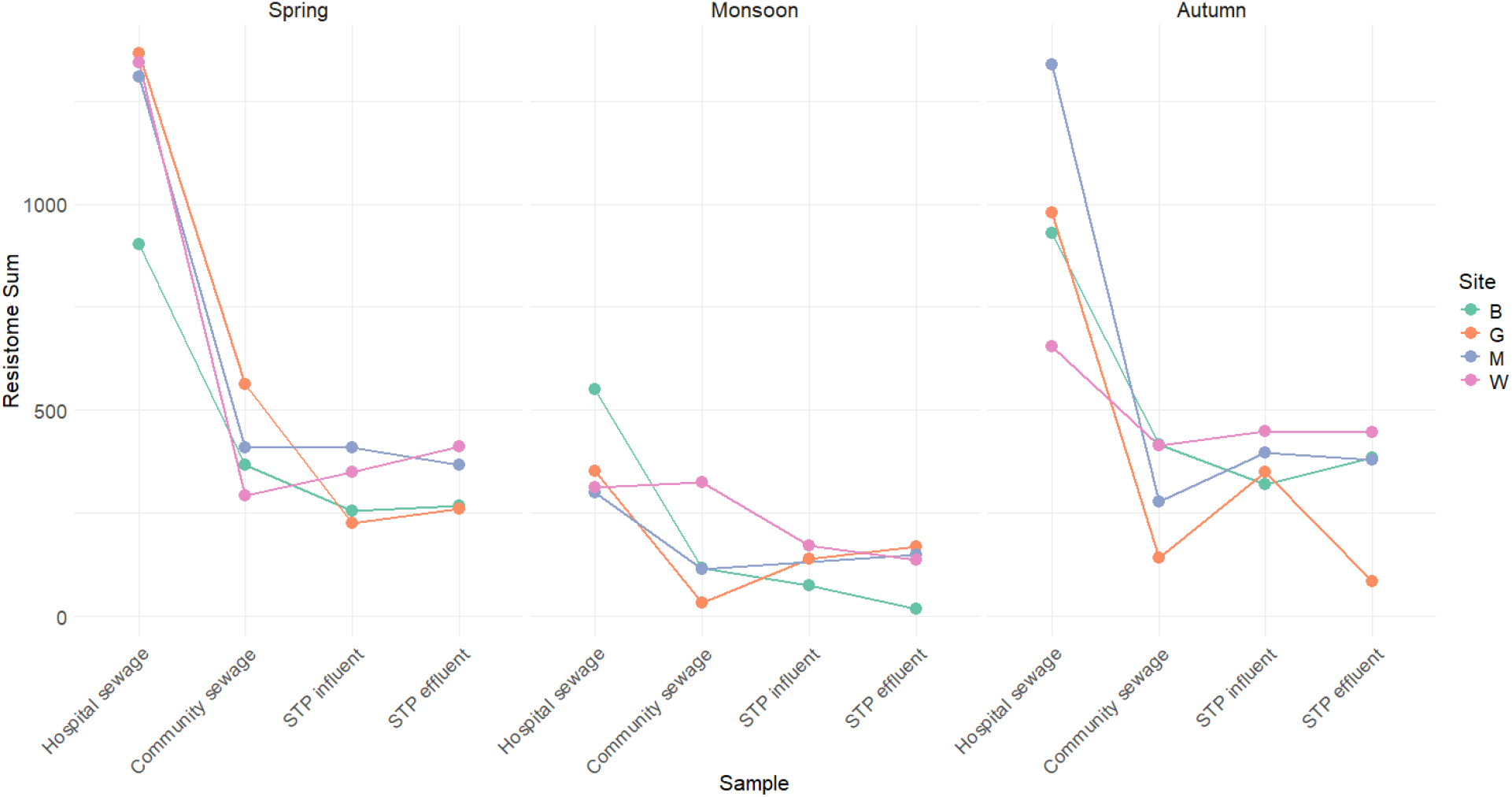
Spatial trends in ARG load. The ARG load (y-axis) is represented by season with sample types along the x-axis and colors indicate the hospital sites.

**Figure 9.**
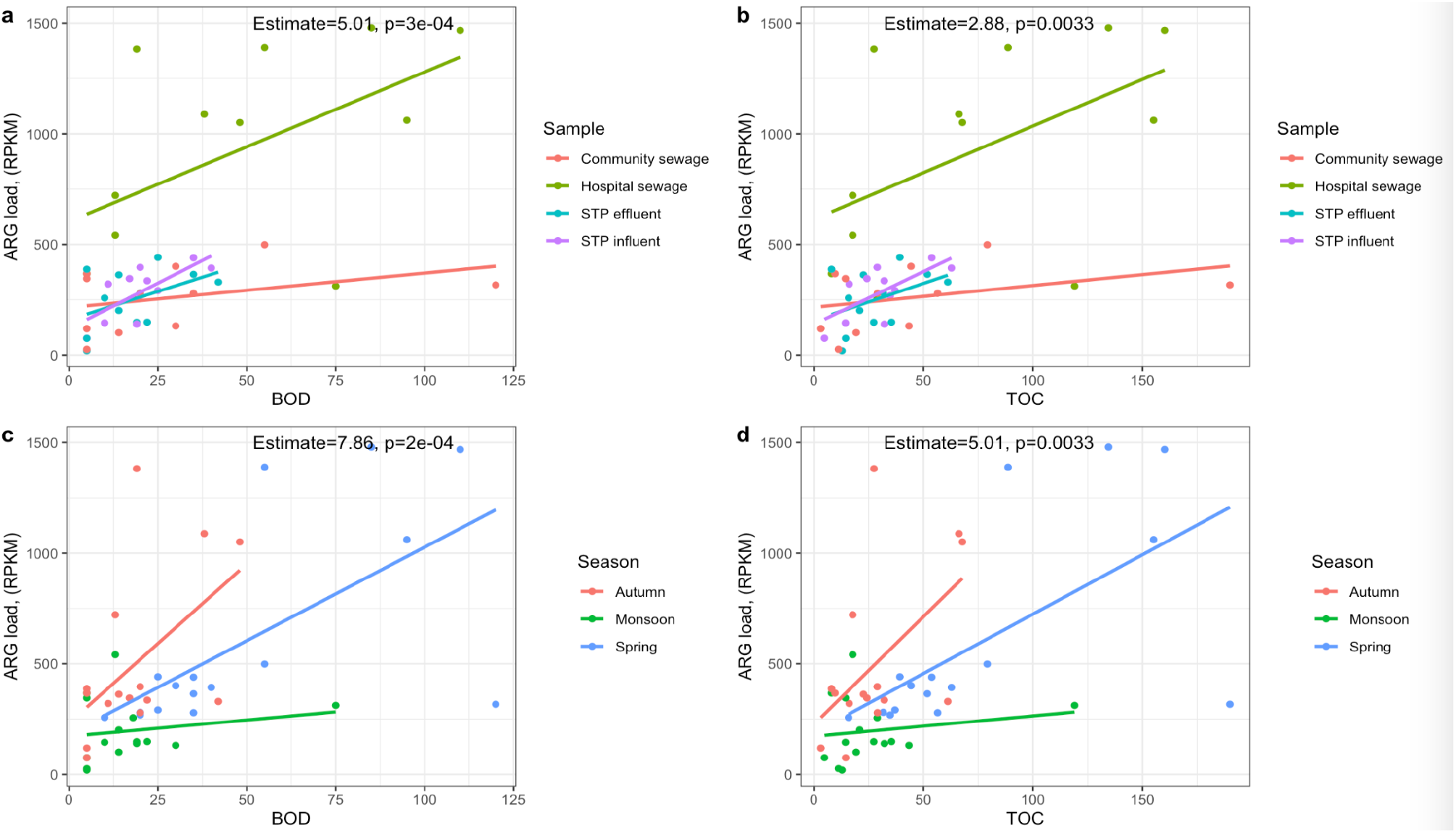
Biological oxygen demand (BOD) and total organic content (TOC) correlate with ARG load. Association between ARG load and **a** BOD and **b** TOC by sampling point (hospital, community, and STP influent and effluent). Association between ARG load and **c** BOD and **d** TOC by season (autumn, monsoon and spring). Estimates and p-values for BOD and TOC from linear models adjusted for sampling points in the network (Sample) or Season are annotated in the respective panels.

## 4. Discussion

We observed a high abundance of bacterial strains with potential clinical significance and ARGs in the sewage treatment plant network. In contrast to our expectation, the ARG load did not exhibit a clear decreasing trend during sewage treatment in our data. The hospital effluent tended to differ from the other points in the sewage network and had the highest relative abundances of potentially pathogenic taxa as well as resistance genes. The beta diversity of the sewage bacterial community and resistome profiles varied by season, with monsoon differing from spring and autumn.

We hypothesized that higher BOD would be associated with higher ARG load, as high nutrient availability favours human-associated bacteria over more slow-growing environmental bacteria (ref). We tested if ARG load was correlated with BOD in the sewage network at the different sampling points and seasons which have differing species and ARG compositions. It was observed that irrespective of season and sampling point in the sewage network, BOD was positively associated with the ARG load. The result suggests that surveilling ARG pollution could be supplemented by using BOD monitoring after the initial ARG load has been quantified and its association with the load established. In our particular case, a unit change in BOD was associated with a change of 10.4 RPKM in the ARG load on average. However, the change in ARG load from influent to effluent was not correlated with the respective change in BOD, which may be due to limited sample size.

The high abundance of potentially pathogenic bacteria and ARGs observed in the sewage networks highlights the urgent need for monitoring the ARG load in sewage. Worryingly, the ARG load or the abundance of potentially pathogenic taxa did not significantly decrease at the STPs, suggesting that sewage treatment could not efficiently replace potentially resistant pathogenic bacteria with bacteria that are typical to the STP microbiome. As a result, hospital and community sewage could potentially represent a major source of pollution in the receiving environments with antibiotic-resistant pathogens. The local conditions in which the average monthly temperatures remain between 20-30^0^C for most of the year can further promote the survival of mesophilic pathogens.

Our study demonstrates that BOD can be used for complementary monitoring of the potential ARG pollution in sewage networks in areas where there is a crucial need for ARG monitoring but scant resources. Previously, faecal pollution measured from shotgun sequences using crAssphage abundance estimation has been shown to track ARG loads (Karkman, et al., 2019). Similarly, tracking the abundance of coliform bacteria can be used to estimate faecal contamination in water. Inexpensive complementary estimation of the ARG pollution could be instrumental in low-resource settings, such as LMICs, with limited availability of molecular biology techniques.

## Data Availability

All data produced in the present work are contained in the manuscript.

## Data and code availability

The text files generated from the metagenomic analysis conducted in this study are available in the GitHub repository (https://github.com/ShivangPB/Wastewater). The custom code used to process these data and perform the analyses is also available in the same repository.

Any supplementary information that supports the findings of this study is available from the corresponding author upon reasonable request.

## Acknowledgements

This research was supported by the Bill & Melinda Gates Foundation under Grant (BT/AMR0308/05/18-NCCS), Alhopuro Foundation(decision number: 20220114) and the Research Council of Finland(decision number:348439 & decision number:333260). The funding agencies played no role in the study design, data collection and analysis, decision to publish, or preparation of the manuscript. The authors wish to acknowledge CSC – IT Center for Science, Finland, for generous computational resources.

## Notes

### Competing Interest Statement

The authors have declared no competing interest.

